# The findings of Antimicrobial Resistance Surveillance in Bangladesh (2016-2020)

**DOI:** 10.1101/2021.06.12.21251710

**Authors:** Zakir Hossain Habib, Saima binte Golam Rasul, Md. Ashraful Alam, Nazmun Nahar Bably, Iqbal Ansary Khan, S.M. Shahriar Rizvi, Tahmina Shirin, Ahmed Nawsher Alam, M. Salim Uzzaman, ASM Alamgir, Mahbubur Rahman, Ashek Ahmmed Shahid Reza, Kazi Mohammad Hassan Ameen, AKM Muraduzzaman, Ishrat Siddiqui, Zarin Tasnim Haider, Meerjady Sabrina Flora

## Abstract

Surveillance is one of the effective tools to address antimicrobial resistance. In Bangladesh a countrywide antimicrobial resistance surveillance has been ongoing since 2016. The main objective of this surveillance is to formulate the guideline for clinicians and to assist policy makers to know the gravity of the AMR problem in Bangladesh.

It is a case-based surveillance conducted by Institute of Epidemiology, Disease Control & Research (IEDCR) in nine sentinel sites where five types of clinical cases were selected according to case definition, and ten types of bacteria were identified from six types of preselected specimens. All the laboratory works were performed following the standard operating procedure supplied by the AMR surveillance Reference laboratory at IEDCR. Total 19,263 samples were processed during the period of March 2017 to March 2020 among which wound swab yielded highest growth (57%). *E. coli* was the highest (1717) isolated organism among the ten priority pathogens which showed highest sensitivity (91%) to Imipenem. Imipenem also showed higher sensitivity to most of the organisms. Third generation cephalosporin was found to be less sensitive to *Escherichia coli* (37%) and *Klebsiella pneumoniae* (28%); nevertheless, *Salmonella* species showed higher sensitivity (97%) to it. *Acinetobacter calcoaceticus-baumannii complex* isolated from ICU patients showed alarming resistance to all of the antibiotics including highest sensitive antibiotic Imipenem (29%). *Salmonella* species isolated from blood showed higher susceptibility to most of the antibiotics except ciprofloxacin (7%). Alarmingly, only 36% of the *Staphylococcus aureus* isolates showed susceptibility to cefoxitin indicates high prevalence of MRSA infection.

The result of the surveillance representing the whole country is surely alarming as many of the bacteria are resistant to the commonly used as well as reserve groups of antibiotics. Concerted effort should be taken from all concerned authorities to curb the problem immediately.

## Introduction

Antimicrobial resistance (AMR) is one of the most complex and multifaceted health challenges facing the global community. It is regarded as the single biggest threat facing the world in the area of infectious diseases. Excessive and inappropriate antibiotic usage is regarded as the main cause of the emergence of resistant organisms. Drug-resistant infections have already contributed to at least 700,000 deaths a year [1]. In Brazil, Indonesia and Russia, 40 to 60% of infections are already caused by drug-resistant bacteria, compared to an average of 17% in Organisation for Economic Co-operation and Development (OECD) countries [2]. Given the current trajectory, drug resistance could lead to 10 million deaths annually and plunge 24 million people into extreme poverty by 2050 [1]. Left unchecked, AMR is likely to become one of the world’s largest health threats, surpassing many other major conditions, such as diabetes and cancer; in scale have a severe effect on economies around the world.

Bangladesh, a developing country of Southeast Asia with a high degree of AMR, poses a regional and global threat. In a study performed in Chittagong in 2003, typhoid patients were found to be unresponsive to second-line therapy (ciprofloxacin). First-line therapy was not even attempted because of existing resistance [3]. Therapeutic failures like this are not rare at all. Furthermore, concerning this, multiple studies have demonstrated irrational antibiotic prescribing by physicians, a habit of self-medication among patients, and the indiscriminate use of antibiotics in agriculture and farming in different parts of the country [4]. Even though many studies have been performed on the prevalence of AMR in Bangladesh, no attempts have yet been made to systematically unify them.

The World Health Organization (WHO) acknowledged AMR as a global public health problem in 1998 and urged member states to take measures to encourage appropriate use of antimicrobials. In May 2015, the World Health Assembly adopted the Global Action Plan on AMR. All countries are required to develop their national action plan based on the GAP. One of the five strategic objectives of the Global Plan is to strengthen the evidence base through surveillance and research. The World Health Organization (WHO) has developed the Global Antimicrobial Resistance Surveillance System (GLASS) to support the implementation of the Global Action Plan on antimicrobial resistance (AMR). GLASS promotes and supports standardized antimicrobial resistance (AMR) surveillance worldwide [1].

In concordance with the global and WHO activities on Antimicrobial Resistance Containment (ARC), the Ministry of Health and Family Welfare (MoHFW) in Bangladesh has come forward and the initiative was taken to conduct the program for containment of antimicrobial resistance in Bangladesh. A National Strategy for ARC in Bangladesh as well as National Action Plan 2017-2022 was developed. The establishment of a Surveillance System for AMR is emphasized in (NAP). A nationwide AMR Surveillance in human health is being conducted by the Institute of Epidemiology, Disease Control & Research (IEDCR) since 2016 keeping in line with GLASS as well as country perspective in nine sentinel sites in different geographical locations all over the country to know the status of Antimicrobial resistance pattern of different bacteria.

## Materials and Methods

The AMR surveillance in Bangladesh is a ‘case-based surveillance’, one of the three surveillance methods designed under the GLASS protocol. With the technical support from the US-CDC through Global Health Security Agenda (GHSA), World Health Organization (WHO) and Government of Bangladesh, IEDCR conducted the surveillance for the period of March 2016 to March 2020.

Surveillance sites were selected on the basis of geographical representation (Figure 2), ability of the hospital to enroll cases and availability of a microbiology laboratory with capacity to perform bacterial culture and sensitivity tests. Capacity building of the sites included hands-on training (both basic & refresher), providing of laboratory standard operating procedures (SOPs), providing of instruments, logistics and technical support through the laboratory networking system.

**Figure 1:**
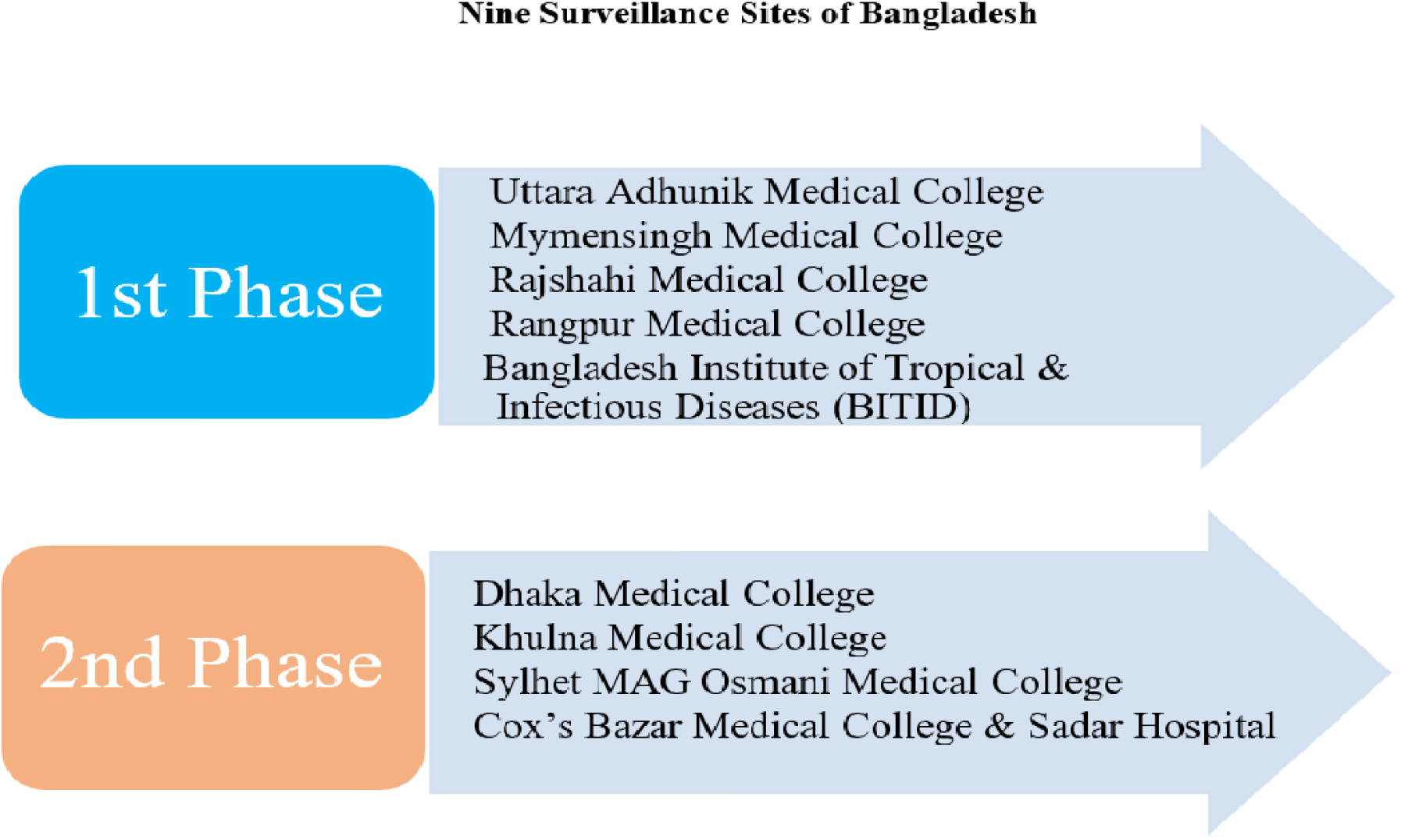
Surveillance activities started in 9 sites all over Bangladesh in 2 phases:

**Figure 2:**
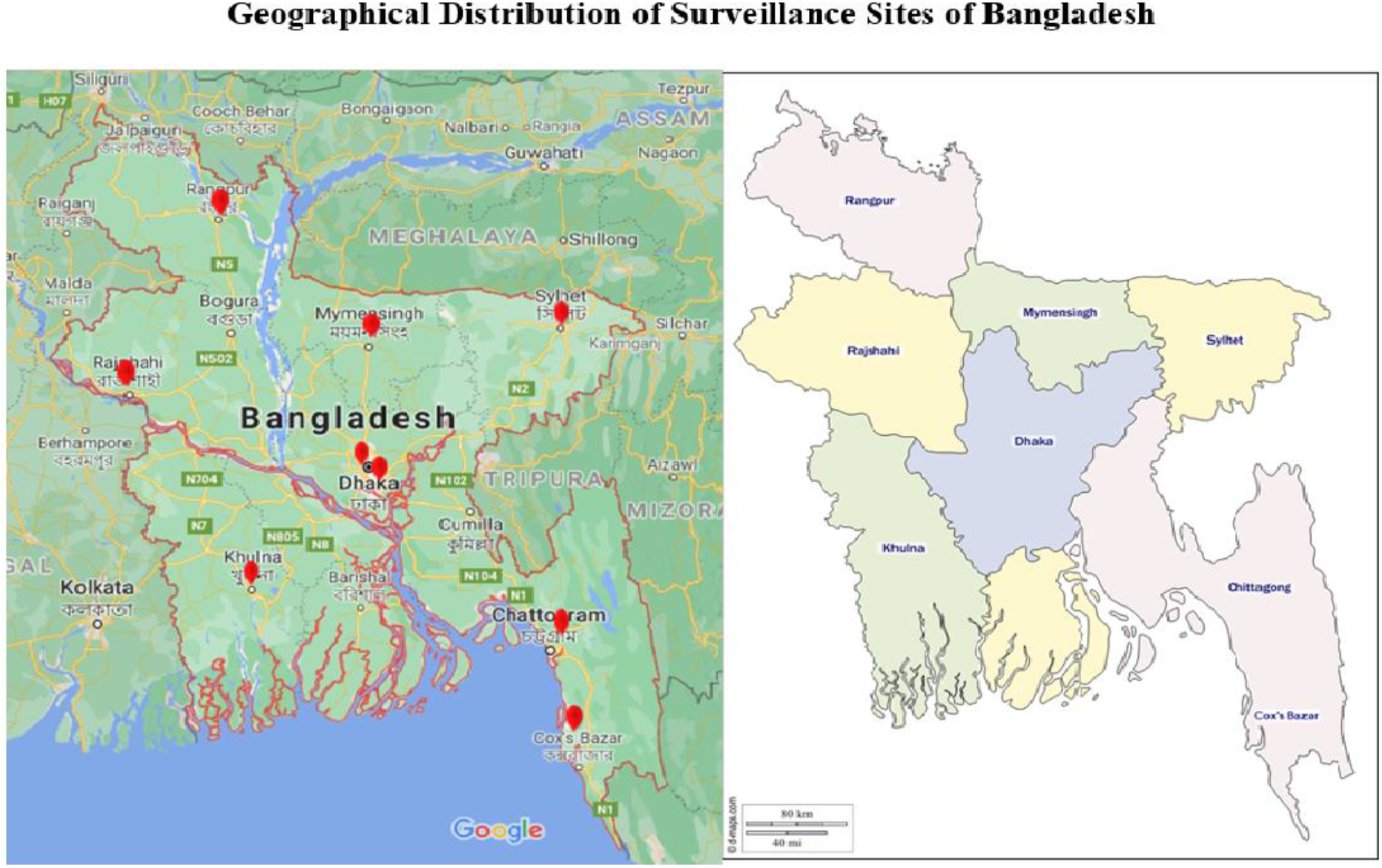
Geographical Distribution of Surveillance Sites of Bangladesh.

Surveillance activities included collection and testing of samples and compiling of the epidemiological and laboratory data. All the laboratory tests including the Antimicrobial Susceptibility Testing (AST) was done at the sentinel sites. Laboratory works have been performed by strictly following the SOPs provided by the AMR Reference Laboratory at IEDCR. All the relevant epidemiological as well as laboratory data were compiled in a software and the hard copy was maintained. The data were cleansed and analyzed using WHONET software at IEDCR.

Internal and external quality control were ensured. For external quality control, 5% of the randomly selected isolates of positive samples had been retested at the reference laboratory as well as discordant results were checked at another quality laboratory. The reference laboratory as well as the sentinel sites participated in Proficiency Testing (PT) by the College of American Pathologists (CAP).

Five different cases of infectious conditions including Urinary tract infection (UTI), diarrhoeal diseases, Wound infection, pneumonia and septicaemia were enrolled from the hospitals by the surveillance physicians following case definitions and specific samples were collected from them according to the protocol which included urine, stool, wound swab, blood, sputum and endotracheal aspirate. In the laboratories, 10 pathogens were identified by biochemical methods from the samples following laboratory SOP and their susceptibility test was done by Kirby-Bauer disc diffusion method following CLSI and also, the zone diameter was noted. The organisms identified were-*Escherichia coli, Klebsiella pneumoniae, Enterococcus* species (*spp*)., *Vibrio cholerae, Shigella spp. Streptococcus pneumoniae, Staphylococcus aureus, Salmonella spp*., *Pseudomonas aeruginosa and Acinetobacter calcoaceticus-baumannii (Acb)* complex. These 10 pathogens were called the priority pathogens.

### Ethical Issue

Patients were selected according to protocol and before taking sample and epi-data informed written as well as verbal consent were taken and other ethical issues were strictly taken into consideration. The protocol was approved by the lnstitutional Review Board (lRB) of lnstitute of Epidemiology Disease Control and Research (IEDCR).

## Result

Figure 3 demonstrates the age & sex distribution according to specimen type.

**Fig 3:**
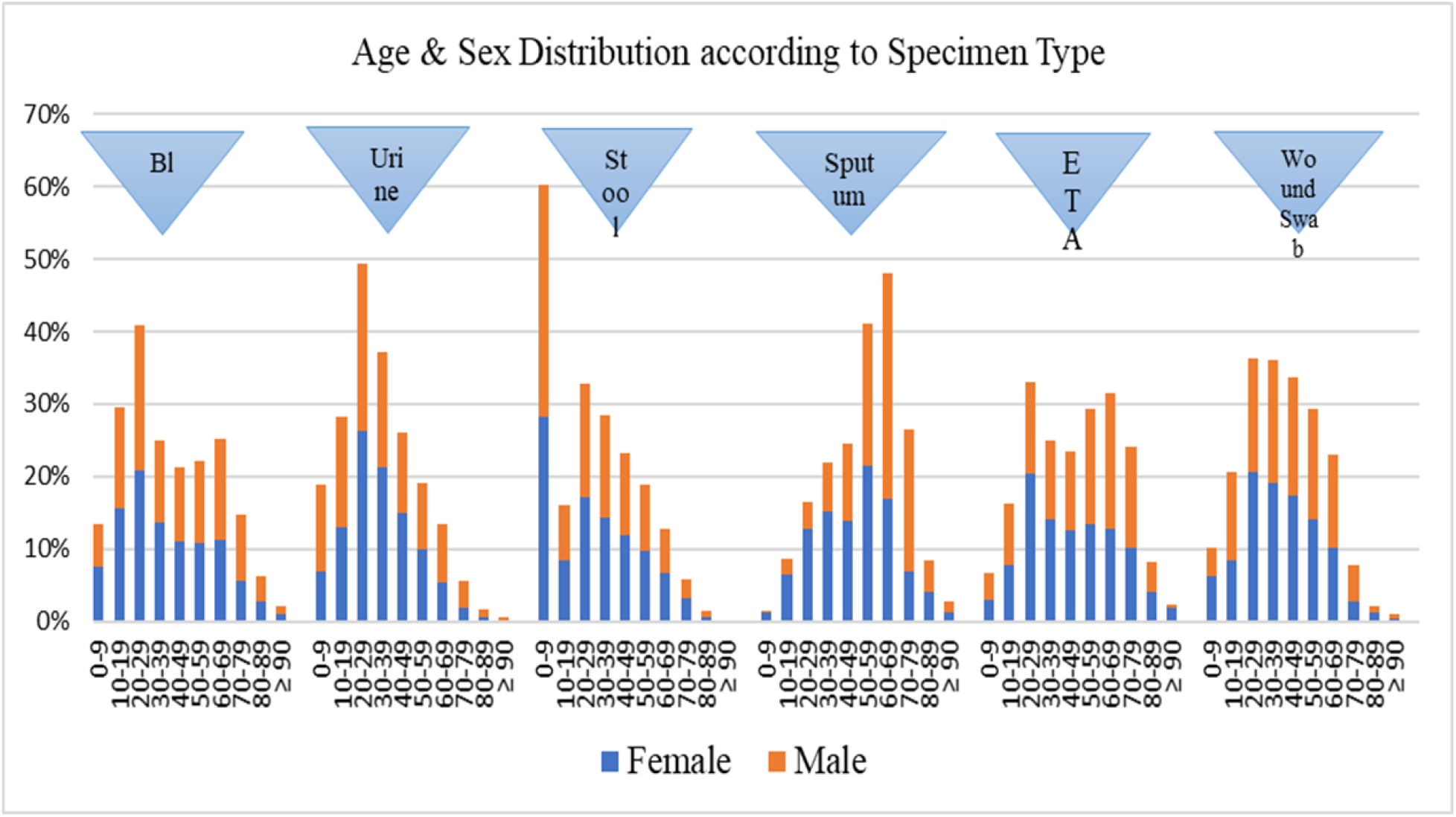
Age & sex distribution according to specimen type

Except stool and sputum, the other 4 samples were mostly collected from the patients between the age group of 10-29 years. The highest number of stool samples were collected from the age group 0-9 years and the highest number of sputum samples were collected from the age group 60-69 years. As per the sex distribution, there was no significance difference between male and female patients.

Figure 4 demonstrates distribution of growth in cultured specimen

**Figure 4.**
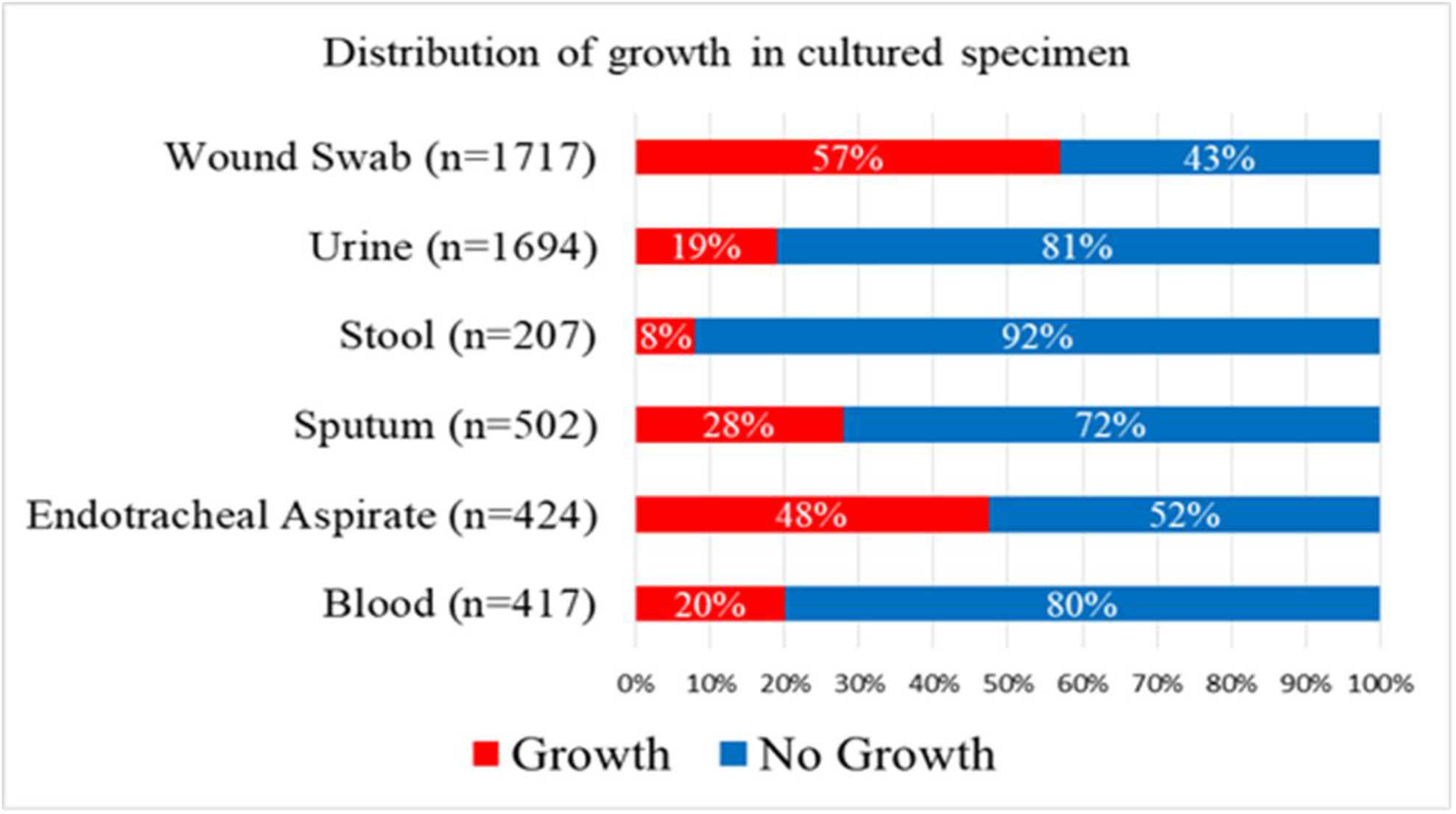
demonstrates distribution of growth in cultured specimen.

Wound swab yielded the highest growth among the 6 different samples (57%) followed by endotracheal aspirate (48%). On the other hand, stool samples yielded the lowest growth (8%). Here, the growth of *E. coli* in stool samples was excluded as the pathogenicity test for the isolates was not done.

Table 1 demonstrates Distribution of priority pathogens according to specimens

**Table 1:** Distribution of priority pathogens according to specimens.

| Priority Pathogens | Specimens |  |  |  |  |  |
| --- | --- | --- | --- | --- | --- | --- |
|  | Wound Swab (n=1717) | Urine (n=1694) | Stool (n=207) | Sputum (n=502) | Endotracheal Aspirate (n=424) | Blood (n=417) |
| *E. coli* | 233(14%) | 1015(60%) | - | 51(10%) | 35(8%) | 65(16%) |
| *Klebsiella pneumoniae* | 156(9%) | 278(16%) | - | 231(46%) | 149(35%) | 31(7%) |
| *Enterococcus species* | 13(0.76%) | 85(5%) | - | 2(0.4%) | - | 1(0.24%) |
| *Vibrio cholerae* | - | - | 110(53%) | - | - | - |
| *Shigella species* | - | - | 47(23%) | - | - | - |
| *Streptococcus pneumoniae* | - | - | - | 2(0.4%) | 1(0.2%) | 3(0.7%) |
| *Staphylococcus aureus* | 235(14%) | 115(7%) | 2(1%) | 95(19%) | 26(6%) | 36(9%) |
| *Salmonella species* | 1(0.06%) | - | 45(22%) | - | - | 243(58%) |
| *Pseudomonas aeruginosa* | 779(45%) | 93(6%) | - | 51(4%) | 80(19%) | 20(5%) |
| *Acb Complex* | 52(3%) | 23(1%) | 1(0.48%) | 10(2%) | 108(26%) | 12(3%) |
| Others | 248(14%) | 85(5%) | 2(1%) | 60(12%) | 25(6%) | 6(1%) |

*Acb, Acinetobacter calcoaceticus-baumannii.*

Among the 10 priority pathogens, *E. coli* showed the highest growth (60%) in the urine samples. Among the wound swabs, *Pseudomonas aeruginosa was the highest isolated organism (45%) followed by Staph. aureus (14%) and E. coli (14%). Vibrio cholera* was the most abundant in stool samples (53%). In the sputum and endotracheal aspirates, *Klebsiella pneumoniae* showed the highest growth (46% and 35% respectively). *Acb* complex was the second highest organism identified from the endotracheal aspirate (26%). In blood, *Salmonella spp*. were the highest isolated (58%) pathogens.

Table 2 demonstrates the antibiotic susceptibility of priority pathogens irrespective of specimen.

**Table 2:** Antibiotic susceptibility of priority pathogens irrespective of specimens.

| Antibiotic Name | Priority Pathogens |  |  |  |  |  |  |  |  |  |
| --- | --- | --- | --- | --- | --- | --- | --- | --- | --- | --- |
|  | *Escherichia coli* (n=1417) | *Klebsiella pneumoniae* (n=845) | *Enterococcus* species (n=101) | *Vibrio cholerae* (n=110) | *Shigella* species (n=47) | *Staphylococcus aureus* (n=509) | *Salmonella* spp. (n=244) | *Non typhoidal Salmonella* (n=45) | *Pseudomonas-aeruginosa* (n=1023) | *Acb Complex* (n=206) |
| **Amikacin** | 80% | 63% | - | - | - | - | 98% | - | 35% | 27% |
| **Amoxicillin-Clavulanate** | 26% | 16% | - | - | - | - | 85% | - | - | - |
| **Ampicillin** | 10% | 3% | 60% | - | 36% | - | 74% | 26% | - | - |
| **Aztreonam** | 34% | 29% | - | - | - | - | 94% | - | 20% | - |
| **Azithromycin** | - | - | - | 83% | 53% | 12% | - | * | - | - |
| **Ceftazidime** | 40% | 29% | - | - | - | - | 98% | - | 20% | 7% |
| **Clindamycin** | - | - | - | - | - | 41% | - | - | - | - |
| **Ciprofloxacin** | 39% | 37% | 38% | 84% | 25% | 30% | 7% | 49% | 23% | 12% |
| **Cephalexin** | 26% | 20% | - | - | - | - | 94% | - | - | - |
| **Cefepime** | 46% | 39% | - | - | - | - | 98% | - | 26% | 13% |
| **Ceftriaxone** | 37% | 28% | - | - | 89% | - | 97% | 70% | - | 9% |
| **Cefoxitin** | - | - | - | - | - | 36% | - | - | - | - |
| **Cefuroxime** | 30% | 25% | - | - | - | - | 95% | - | - | - |
| **Doxycycline** | - | - | - | - | - | 66% | - | - | - | 27% |
| **Erythromycin** | - | - | - | 7% | - | - | - | - | - | - |
| **Gentamicin** | 70% | 57% | 40% | - | - | 63% | 98% | - | 25% | 20% |
| **Imipenem** | 91% | 77% | - | - | - | - | 100% | - | 53% | 29% |
| **Linezolid** | - | - | 76% | - | - | - | - | - | - | - |
| **Mecillinam** | - | - | - | - | 64% | - | - | - | - | - |
| **Nitrofurantoin** | 77% | 46% | 75% | - | - | - | - | - | - | - |
| **Norfloxacin** | 45% | 49% | - | - | - | - | 79% | - | 18% | - |
| **Oxacillin** | - | - | - | - | - | 21% | - | - | - | - |
| **Piperacillin** | 22% | 17% | - | - | - | - | 74% | - | 30% | 6% |
| **Piperacillin-Tazobactam** | - | - | - | - | - | - | - | - | 38% | 20% |
| **Penicillin-G** | - | - | 38% | - | - | 11% | - | - | - | - |
| **Rifampicin** | - | - | - | - | - | 69% | - | - | - | - |
| **Sulfamethoxazole-Trimethoprim** | 41% | 31% | - | 4% | 37% | 49% | 82% | 51% | - | 21% |
| **Tetracycline** | 42% | 37% | 26% | 57% | - | - | 85% | - | - | - |

\*Less than 30 isolates tested.

A varied range of antibiotic susceptibility is found in different bacteria. Most of the bacteria were susceptible to Imipenem; among them *Salmonella* was 100% susceptible. Ampicillin was less effective against pathogens like *E. coli, Klebsiella pneumoniae* and Non-typhoidal *Salmonella. Salmonella* spp. showed high susceptibility to almost all antibiotics. Among the 10 priority pathogens, *Pseudomonas aeruginosa* and *Acb complex* showed low sensitivity to almost all the antibiotics. *Pseudomonas aeruginosa* showed less than 40% sensitivity to all the antibiotics except imipenem and *Acb complex* showed less than 30% sensitivity to all of the antibiotics. Only 36% *Staphylococcus aureus* isolates were susceptible to Cefoxitin whereas Clindamycin was sensitive to 41% isolates.

The susceptibility patterns of *Staphylococcus aureus* to vancomycin and Linezolid are not mentioned here as those could not be confirmed by MIC testing.

Table 3 demonstrates antibiogram of urine sample

**Table: 3.** Antibiogram of Urine.

| **Antibiotic Name** | ***Escherichia coli***<br>**(n=1015)** | ***Klebsiella pneumoniae***<br>**(n=278)** | ***Enterococcus* species**<br>**(n=85)** | ***Staphylococcus aureus***<br>**(n=115)** | ***Pseudomonas aeruginosa***<br>**(n=93)** |
| --- | --- | --- | --- | --- | --- |
| **Amikacin** | 86% | 82% | - | - | 69% |
| **Amoxicillin-Clavulanate** | 30% | 31% | - | - | - |
| **Ampicillin** | 12% | 5% | 61% | - | - |
| **Aztreonam** | 39% | 51% | - | - | 30% |
| **Azithromycin** | - | - | - | 7% | - |
| **Ceftazidime** | 45% | 54% | - | - | 36% |
| **Clindamycin** | - | - | - | 45% | - |
| **Ciprofloxacin** | 44% | 57% | 41% | 41% | 40% |
| **Cephalexin** | 29% | 36% | - | - | - |
| **Cefepime** | 52% | 63% | - | - | 44% |
| **Ceftriaxone** | 45% | 55% | - | - | - |
| **Cefoxitin** | - | - | - | 20% | - |
| **Cefuroxime** | 35% | 43% | - | - | - |
| **Doxycycline** | - | - | - | 70% | - |
| **Gentamicin** | 78% | 75% | 46% | 75% | 63% |
| **Imipenem** | 95% | 92% | - | - | 74% |
| **Linezolid** | - | - | 80% | - | - |
| **Nitrofurantoin** | 80% | 51% | 75% | - | - |
| **Norfloxacin** | 46% | 59% | - | - | * |
| **Oxacillin** | - | - | - | 7% | - |
| **Piperacillin** | 25% | 32% | - | - | 53% |
| **Piperacillin-Tazobactam** | - | - | - | - | 73% |
| **Penicillin-G** | - | - | 37% | 24% | - |
| **Rifampicin** | - | - | - | 83% | - |
| **Sulfamethoxazole-Trimethoprim** | 45% | 45% | - | 53% | - |
| **Tetracycline** | 45% | 49% | 26% | - | - |

\*Less than 30 isolates tested.

Five types of pathogens were identified from the urine sample, where *E. coli* was highest, followed by *Klebsiella pneumoniae*. Imipenem was the most effective against the pathogens identified from the urine samples. More than 90% of isolates of *E. coli* and *Klebsiella pneumoniae* were sensitive to Imipenem whereas ampicillin showed lowest sensitivity to both of these organisms (12% and 5% respectively). Only 45% of the *E. coli* were susceptible to Ceftriaxone. Nitrofurantoin was sensitive to 80% of *E. coli*. The antibiotics showed relatively less sensitivity to *S. aureus* and *P. aeruginosa* pathogens. About 20% and 45% of the *S. aureus* isolates were sensitive to Cefoxitin and clindamycin respectively.

Table 4 demonstrates the antibiogram of the wound swab samples.

**Table: 4.**
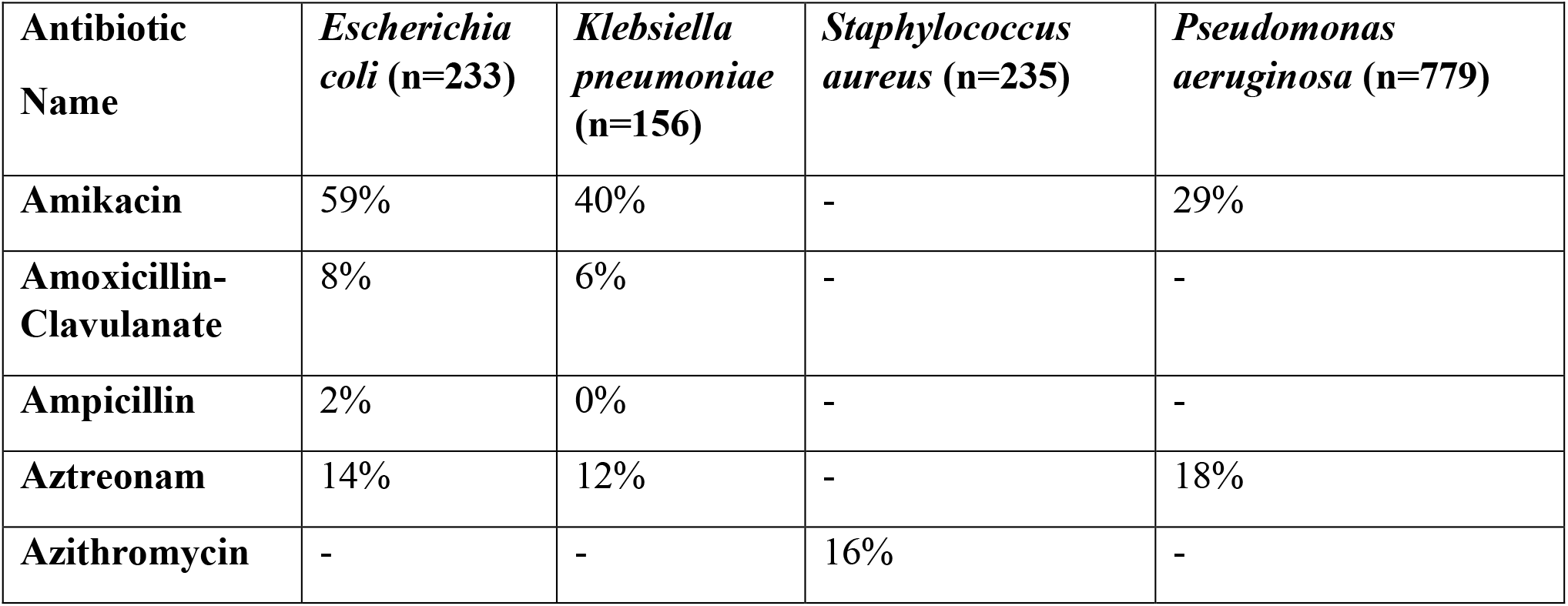
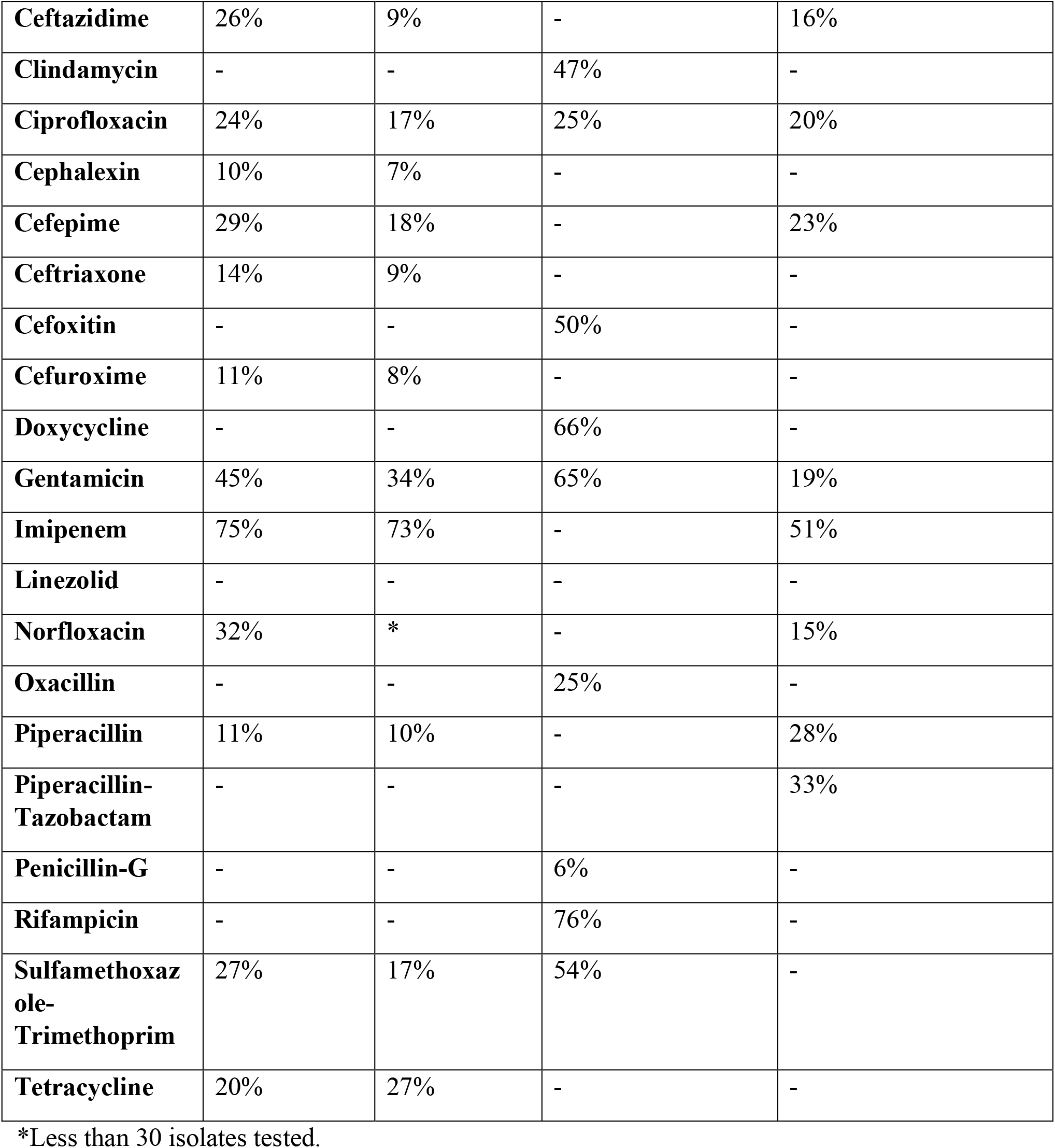
Antibiogram of Wound Swab.

Both *E. coli* and *K. pneumoniae* showed highest susceptibility (75% and 73% respectively) to Imipenem. Ceftriaxone showed sensitivity to only 14% and 9% of *E. coli* and *K. pneumoniae* isolates respectively. Other than Imipenem, other antibiotics were sensitive to less than 35% of *P. aeruginosa*. Most *S. aureus* (76%) were susceptible to Rifampicin whereas fewest (6%) were susceptible to Penicillin-G. Half of the *S. aureus* isolates were susceptible to cefoxitin and 47% of the isolates showed susceptibility to clindamycin.

Table 5 demonstrates the antibiogram of the Stool sample.

**Table: 5.** Antibiogram of Stool.

| **Antibiotic Name** | ***Vibrio cholerae***<br>**(n=110)** | ***Shigella* species**<br>**(n=47)** | ***Salmonella***<br>**species (n=45)** |
| --- | --- | --- | --- |
| **Ampicillin** | - | 36% | 26% |
| **Azithromycin** | 83% | * | * |
| **Ciprofloxacin** | 84% | 25% | 49% |
| **Ceftriaxone** | - | 89% | 70% |
| **Erythromycin** | 7% | - | - |
| **Mecillinam** | - | 64% | - |
| **Sulfamethoxazole-Trimethoprim** | 4% | 37% | 51% |
| **Tetracycline** | 57% | - | - |

\*Less than 30 isolates tested.

Ceftriaxone showed the highest sensitivity to *Shigella* and *Salmonella* species (89% and 70% respectively). Ciprofloxacin showed the highest sensitivity (84%) to *V. cholerae*, followed by Azithromycin (83%), their sensitivity being almost similar.

Table 6 demonstrates the antibiogram of the sputum sample.

**Table: 6.**
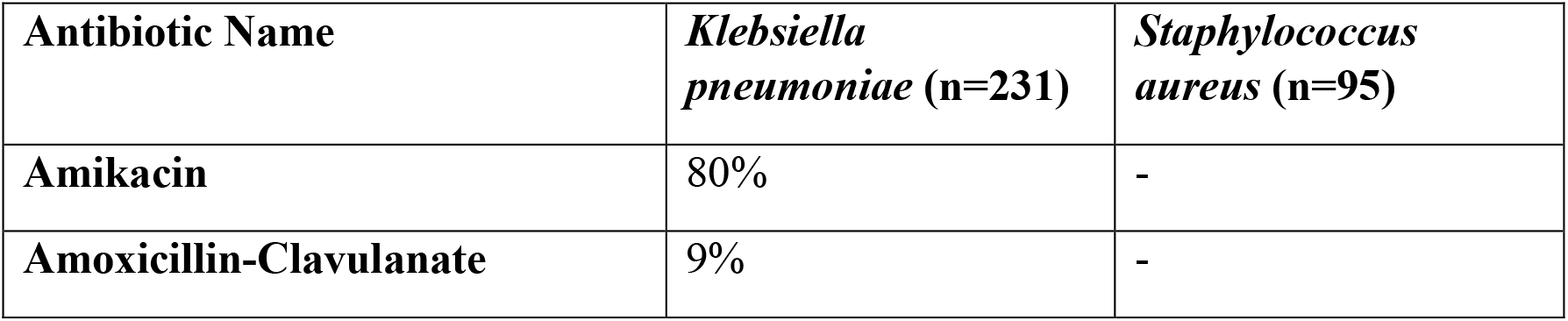
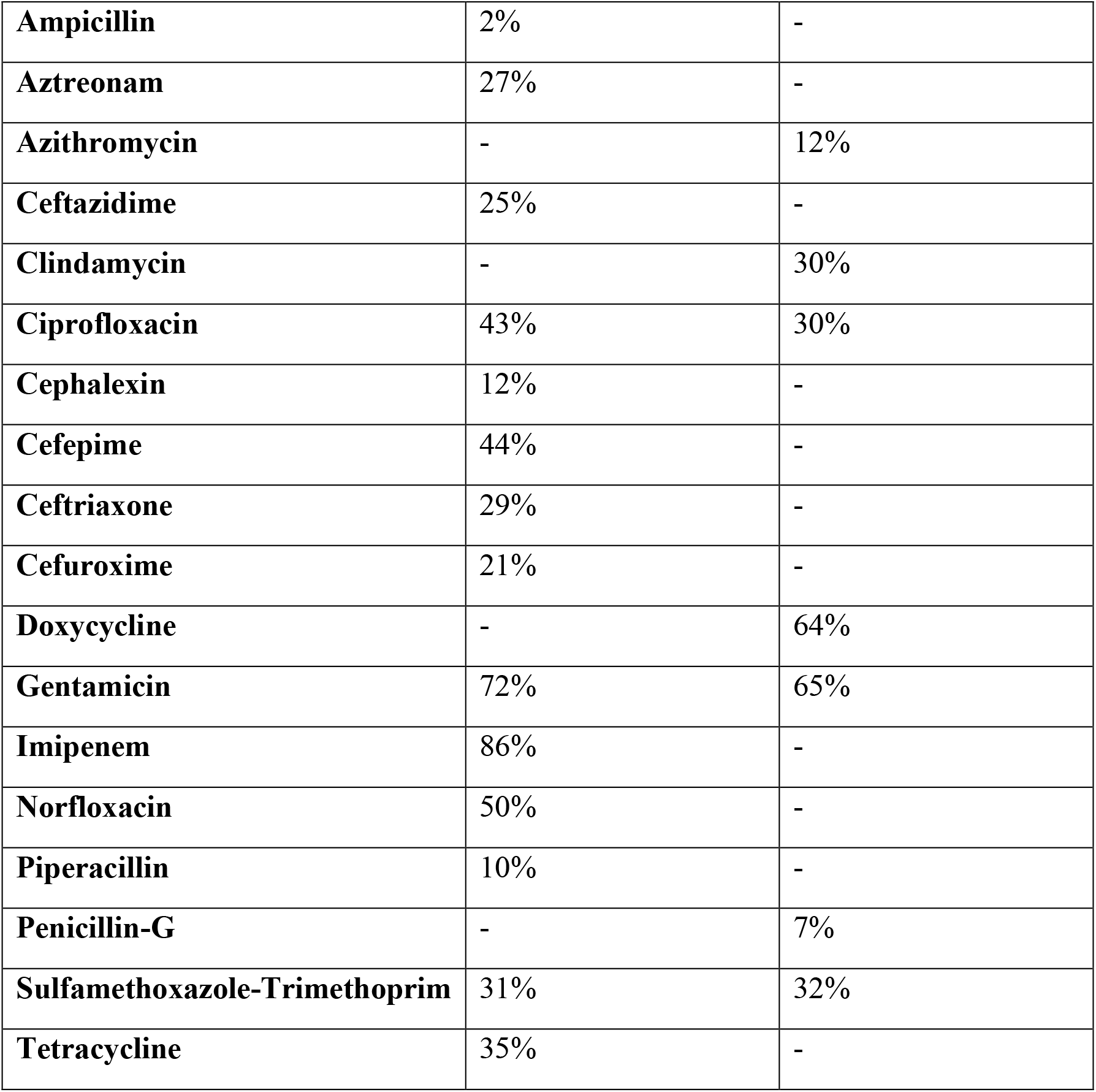
Antibiogram of Sputum.

Imipenem showed the highest sensitivity to *K. pneumoniae* (86%), followed by Amikacin (80%). Gentamicin showed the highest sensitivity to S. aureus. *S. pneumoniae* was not included in the table as the number of the isolates was less than 30.

Table 7 demonstrates the antibiogram of endotracheal aspirate pathogens

**Table: 7.** Antibiogram of ETA.

| **Antibiotic Name** | ***Klebsiella pneumoniae* (n=149)** | ***Pseudomonas aeruginosa* (n=80)** | ***Acb* Complex (n=108)** |
| --- | --- | --- | --- |
| **Amikacin** | 28% | 25% | 16% |
| **Amoxicillin-Clavulanate** | 5% | - | - |
| **Ampicillin** | 0% | - | - |
| **Aztreonam** | 3% | 10% | - |
| **Ceftazidime** | 4% | 5% | 2% |
| **Ciprofloxacin** | 9% | 12% | 4% |
| **Cephalexin** | 0% | - | - |
| **Cefepime** | 7% | 6% | 3% |
| **Ceftriaxone** | 2% | - | 4% |
| **Cefuroxime** | 1% | - | - |
| **Doxycycline** | - | - | 28% |
| **Gentamicin** | 24% | 13% | 7% |
| **Imipenem** | 37% | 25% | 15% |
| **Piperacillin** | 1% | 14% | 3% |
| **Piperacillin-Tazobactam** | - | * | 4% |
| **Sulfamethoxazole-Trimethoprim** | 12% | - | 15% |
| **Tetracycline** | 14% | - | - |

\*Less than 30 isolates tested.

Endotracheal aspirate was taken from the ICU patients and all the antibiotics showed low sensitivity to the identified pathogens from this sample (less than 40%). Imipenem was sensitive to only 37% *K. pneumoniae*, which was the highest and only to 25% of *P. aeruginosa*. Amikacin was sensitive to 28% of *K. pneumoniae* and 25% of *P. aeruginosa*. Only 20% or less *Acb* complex isolates were sensitive to all antibiotics. Cefepime, a Reserve drug, showed sensitivity to less than 10% of all 3 pathogens.

Table 8 demonstrates the antibiogram of blood samples.

**Table: 8.** Antibiogram of Blood.

| **Antibiotic Name** | ***E. coli* (n=65)** | ***Klebsiella pneumoniae* (n=31)** | ***Staphylococcus aureus* (n=36)** | ***Salmonella* species (n=243)** |
| --- | --- | --- | --- | --- |
| **Amikacin** | 74% | 50% | - | 98% |
| **Amoxicillin-Clavulanate** | 27% | * | - | 85% |
| **Ampicillin** | 9% | * | - | 74% |
| **Aztreonam** | 24% | * | - | 94% |
| **Azithromycin** | - | - | 13% | - |
| **Ceftazidime** | 30% | 33% | - | 98% |
| **Clindamycin** | - | - | 35% | - |
| **Ciprofloxacin** | 32% | 27% | 42% | 7% |
| **Cephalexin** | 17% | * | * | 94% |
| **Cefepime** | 32% | * | - | 98% |
| **Ceftriaxone** | 18% | * | - | 97% |
| **Cefuroxime** | 19% | * | - | 95% |
| **Doxycycline** | - | - | 80% | - |
| **Gentamicin** | 63% | * | 57% | 98% |
| **Imipenem** | 89% | * | - | 100% |
| **Norfloxacin** | 15% | * | - | 82% |
| **Piperacillin** | 14% | * | - | 74% |
| **Sulfamethoxazole-Trimethoprim** | 25% | 23% | * | 82% |
| **Tetracycline** | 46% | * | - | 86% |

\*Less than 30 isolates tested.

Among the identified pathogens from the blood sample, *Salmonella* was found to be most abundant. Almost all the antibiotics other than Ciprofloxacin were sensitive to 70% and above of the all *Salmonella* species. Ciprofloxacin showed sensitivity to 7% isolates of the *Salmonella* only. Imipenem was found to be sensitive to all *Salmonella* isolates. *E. coli was found* to be highly susceptible to Imipenem (89%). Eighty percent of S. aureus were susceptible to Doxycycline. Almost 50% or more of all identified pathogens were susceptible to Amikacin.

## Discussion

Like other LMICs, Bangladesh is also facing the problem of AMR and the extent of this AMR is not clear due to the lack of adequate data. The AMR surveillance in Bangladesh is the first of its kind which covered all geographical areas in Bangladesh to find out the resistance pattern of ten important bacterial pathogens from six types of samples collected from patients attending outdoors and indoors presented with five clinical syndromes.

Total 19,263 samples were processed. Other than stool and sputum, most of the samples were collected from the age group of 10-29 years. Stool sample was mostly taken from the pediatric department (the age group was between 0 and 9 years). Some of the stool samples were also collected from the Infectious Disease Hospital and ORT unit of Medical college hospitals. Sputum was mostly collected from the age group of 60-69 years. The gender distribution showed no significant variation in terms of collection of samples.

Even though among the collected samples, urine was the highest in number, it yielded only 19% growth. A study done in a tertiary hospital in Bangladesh from outdoor and indoor samples yielded similar results where 20.1% showed significant growth of bacteria (5).

The lowest growth in culture was yielded by stool (9%). This may be due to exclusion of growth of *E. coli* as its pathogenicity test could not be done. Wound swab and ETA yielded the highest growth, the percentage being 57% and 48% respectively.

Among the gram negative pathogens, *E. coli* showed the highest (28%) growth followed by *Pseudomonas aeruginosa* (21%), *Klebsiella pneumonia* (13%), *Salmonella* species (6%), *Acb complex* (4%), *Vibrio cholera* (2%) and *Shigella* species (1%).

*E*.*coli* yielded highest growth (60%) in urine which is in line with other studies like one conducted in a tertiary care hospital in Bangladesh where *E. coli* (58.18%) was the most prevalent bacteria isolated from positive urine samples (6) and also *E. coli* showed mostly resistant to penicillins and cephalosporins. Protein synthesis inhibitors also showed poor susceptibility patterns. In UTI, nitrofurantoin is the only oral drug that showed a better (80%) susceptibility profile. Imipenem was the most effective against *E. coli* followed by Amikacin. *E. coli* isolated from the wound swab showed a more resistant profile than other samples. In Indian surveillance system, they found colistin, imipenem, meropenem, amikacin and gentamicin as effective drugs which have a concordance with the present study (7). They also found combinations like piperacillin-tazobactam and cefoperazone-sulbactam as effective drugs, which could not be evaluated in our setup. The Indian studies found ESBL producing *E. coli* were responsible for such resistance patterns for penicillin and cephalosporin (8, 9).

*Pseudomonas* species showed the second highest growth following *E. coli*. It is the highest isolated pathogen in wound swab (45%). The samples were collected mainly from the surgery ward followed by the burn unit and medicine unit. A study in Nigeria showed similar result where *Pseudomonas aeruginosa* was the highest (33.3%) isolated pathogen in post-operative patients followed by *Staphylococcus aureus* (21.7%) (10). While a tertiary care hospital in Nepal, pus and wound swab samples from paediatric patients *S. aureus* was the highest isolated organism followed by *P. aeruginosa* (11).

The overall susceptibility pattern of *Pseudomonas* was very poor. *Pseudomonas* were highest susceptible (53%) to Imipenem followed by Piperacillin-Tazobactam (38%) and Amikacin (35%). Only Piperacillin showed 38% sensitivity to *Pseudomonas*. All other antibiotics showed poor sensitivity to *Pseudomonas*. Isolates of *Pseudomonas aeruginosa* from ETA were most resistant. Studies across India showed better susceptibility patterns with colistin followed by aminoglycosides (amikacin/ gentamicin), piperacillin/tazobactam, cephalosporins, fluoroquinolones and carbapenems (imipenem/ meropenem) (7, 12, 13). Multidrug resistance pattern of *Pseudomonas aeruginosa* has already been established (14).

*K. pneumoniae* mostly grew from sputum (46%) and ETA (35%). A study done in Nigeria also found *K. pneumoniae* as the highest isolated pathogen followed by *H. influenzae* and then *S. aureu*s (15). Another study done in a tertiary care hospital, Bali, Indonesia found *K. pneumoniae* as the highest isolated pathogen followed by *Acinetobacter baumannii* (16).

In this study ETA was cultured from the ventilated patient’s samples at ICU and *K. pneumoniae* was found to be the highest pathogen followed by *Acb complex*. Most of the bacteria isolated were gram negative bacilli (86%). Among the gram positive bacteria, *S. aureus* was the highest one (6%). A prospective study done in a tertiary care hospital in Pondicherry, India showed similar results: most cases of Ventilator Associated Pneumonia (VAP) were caused by Gram negative bacteria, which accounted for 80.9% of the causative organisms. *Acinetobacter baumannii* (23.4%) and *Pseudomonas aeruginosa* (21.3%) were the predominant Gram-negative bacteria associated with VAP, and *Staphylococcus aureus* (14.9%) was the most common Gram-positive bacterium among patients with VAP (17).

The overall susceptibility pattern of *K. pneumoniae* showed below 50% for most of the antibiotics except Imipenem (77%), Amikacin (63%) and gentamicin (57%). Isolates from ETA showed the most resistant profile than others. Susceptibility showed diversity in India with high rates of ESBL production. Susceptibility to carbapenems showed a wide range from 44 to 72%, amikacin showed 65% while gentamicin susceptibility was 55% (18, 19).

More than half (58%) of the isolates from blood were *Salmonella* species. Isolates from blood showed an excellent sensitivity pattern except ciprofloxacin (7%) and ampicillin (74%). In India, *Salmonella enterica serovar Typhi* and *Paratyphi* were predominant blood isolates. In *S. Typhi* and *paratyphi*, they found nalidixic acid as the most resistant drug with a variable finding of ciprofloxacin (0-81%). Other drugs showed better sensitivity. Stool constituted 20% of *Salmonella* species. Isolates from stool samples here showed a very different scenario with a highest sensitivity (70%) to ceftriaxone assuming that they might be non typhoidal salmonellosis (NTS). Among NTS from India, susceptibility was observed for ampicillin (0-93%), co-trimoxazole (42-93%), chloramphenicol (45-100%), nalidixic acid (23-76%), ciprofloxacin (9-100%), ceftriaxone and azithromycin (>90%) (20-23). *Shigella* species from stools also have similar sensitivity patterns.

*V. Cholera* was the most abundant bacteria (48%) in stool. Two of the major causes of severe diarrhea in low-resource countries, are the bacterial pathogens *Vibrio cholerae* (O1), which causes epidemic cholera, and enterotoxigenic Escherichia coli (ETEC) (24). We excluded *E. coli* as there is a lack of capacity to toxigenicity in our laboratory set up. In a systemic surveillance on diarrheal patients carried out by icddr, b, it was found that the most prevalent pathogen isolated was *Vibrio cholerae* O1 (23%) followed by ETEC (11%) (25).

In the present study, *Vibrio cholerae* was highly susceptible to ciprofloxacin and azithromycin. In India, the antimicrobial susceptibility profile observed were, ampicillin (0-68%), trimethoprim-sulfamethoxazole (0-33%), chloramphenicol (30-90%), tetracycline (2-70%), nalidixic acid (0-66%), ciprofloxacin (10-94%), norfloxacin (14-94%), ofloxacin (6-90%) and cefixime (0-95%) (26-28).

Though *Acinetobacter baumannii (ACB) complex* comprises only 4% of total and constituted 26% of ETA isolates. Susceptibility profile was the worst as expected and showed the most sensitivity to Imipenem by 29%. But studies from India got better susceptibility patterns for imipenem, meropenem, amikacin, tobramycin, netilmicin and colistin compared to our surveillance reports (29-31).

Among the gram positive organisms, *Staphylococcus aureus* was the most (10%) prevalent, followed by *Enterococcus* species and *Streptococcus pneumoniae. Staphylococcus aureus* isolates were mostly found in respiratory samples followed by wound swab, blood and urine. The overall susceptibility pattern showed the most effective drug was Rifampicin (69%) and the least was Penicillin-G (11%). Only 36% isolates showed sensitivity towards Cefoxitin indicates the high MRSA burden in the region. It still needs to differentiate the healthcare associated and community acquired infections to identify the actual magnitude of the MRSA in these settings. We could not evaluate the source of the infection either hospital origin or community origin. The overall clindamycin susceptibility was higher (41%) than the cefoxitin albeit we could not evaluate the clinical significance of these findings due to lack of performing the inducible clindamycin resistance test in our sentinel sites. In India, *S. aureus* (MSSA) and methicillin-resistant *S. aureus* (MRSA) were reported separately. They found almost 40% isolates as MRSA. For MRSA, poor susceptibility to gentamicin (28-44%), erythromycin (9-69%), clindamycin (35-71%), co-trimoxazole (27-66%) and ciprofloxacin (8-21%) was found (32-34).

*Enterococcus* isolates were mostly present in the Urine. The most sensitive drug was Nitrofurantoin to be found effective against *Enterococcus*. Susceptibility patterns in Indian studies were ampicillin (3-35%), gentamicin (16-89%), vancomycin (77-100%) and linezolid (98-100%). Vancomycin resistance was reported around 20% (35-37). Only 06 *Streptococcus pneumoniae* could be isolated from respiratory samples and blood. The fastidious nature of this organism could be underestimated. Very few isolates are not conclusive to evaluate the antibiogram as it was below 30 in number.

### Limitations of the study

As the participatory laboratories were not capable of performing MIC testing, some of the important susceptibility patterns could not be confirmed which required it according to CLSI. Furthermore, the toxigenicity test of stool samples yielding *E*.*coli* could not be performed so that one of the important diarrheagenic pathogens had to be excluded from the susceptibility list.

## Conclusion

AMR Surveillance is an essential tool for getting necessary information to develop and monitor therapy guidelines, antibiotic stewardship programmes, public health interventions and infection control policies. It enables early detection of resistant strains of public health importance, and supports the prompt notification and investigation of outbreaks and ultimately guides to policy recommendations.

Running a standard and reliable surveillance is extremely important as well as challenging especially in low resource countries like Bangladesh. The ongoing AMR surveillance program, with all its limitations and challenges, is unique for its countrywide expansion and coordinated approach of hospital physicians, nurses of medical college hospitals and laboratory personnel of microbiology departments of the medical colleges. The susceptibility pattern of different microorganisms as revealed by the surveillance is indeed alarming. Except Imipenem most of the commonly used antibiotics showed ineffective for most of the bacteria. The *Acb* complex, most of which organism has been found from ICU patient’s specimens, showed less than 50% sensitivity to all of the used antibiotics. Vibrio cholerae isolated from stool samples showed similar sensitivity to ciprofloxacin (84%) and azithromycin (83%) whereas Shigella spp. showed only 53% sensitivity to azithromycin while ceftriaxone (89%) showed highest sensitivity. Salmonella spp. from blood showed high sensitivity (70-100%) to most of the antibiotics except ciprofloxacin (7%). *Staph. aureus* is only 36% sensitive to cefoxitin which indicates the possibility to a very high number of MRSA. Antibiotics like ampicillin, sulfamethoxazole-trimethoprim and tetracycline which was previously used commonly does not seem to be effective to most of the bacteria except salmonella spp. It is high time all relevant stakeholders should come forward to curb the upcoming threat.

## Data Availability

All the data are obtained from patients culture and sensitivity reports. Demographic data are obtained as the protocol of AMR surveillance in Bangladesh

## Acknowledgement

The AMR surveillance is funded by the Government of People’s Republic of Bangladesh, Centers for Disease Control and Prevention, USA through Global Health Security Agenda and World Health Organizations. The protocol of this surveillance system was developed with the support of American Society for Microbiology. We also acknowledge all the staff of National Reference Laboratory of AMR, IEDCR, Bangladesh and staff of the sentinel sites for their intense support for this study.

## Notes

### Competing Interest Statement

The authors have declared no competing interest.

### Author Declarations

Patients were selected according to national AMR surveillance protocol and before taking sample and epi-data informed written as well as verbal consent were taken and other ethical issues are strictly taken into consideration. The protocol was approved by the lnstitutional Review Board (lRB) of lnstitute of Epidemiology Disease Control and Research (IEDCR).

